# Antibiotic susceptibility trend estimation for India based on 2018 to 2021 data from GEARS program

**DOI:** 10.1101/2024.09.11.24313502

**Authors:** Roshan Ratnakar Naik, Nehal Kalita, Rajesh Kumar Maurya, Annie Rajan

**Author notes:** Corresponding author: Roshan Ratnakar Naik.

## Abstract

Antimicrobial resistance (AMR) continues to be a major public threat due to dwindling supplies of effective antibiotics due to its excessive usage in humans and food producing animals. The objective of our study was to identify and predict future trends in multi-drug resistance amongst enterobacteriaceae and non-enterobacteriaceae family in India for four common broad-spectrum antibiotics. We focussed on four broad spectrum antibiotics levofloxacin, gentamicin, cefepime, and ceftazidime and classified the GEARS program study dataset into sensitive (S), intermediate(I) and resistant (R) isolates based on CLSI breakpoints. Levofloxacin (98.3%) was found to be the most susceptible broad spectrum antibiotic for treatment of non-enterobacteriaceae while it was relatively resistant (56.5%) for enterobacteriaceae treatment. As levofloxacin are among ICMR list of ‘alert antimicrobial agents’,hospitals and clinicans can exercise greater care before prescribing them.

## Introduction

The discovery of antibiotics in the last century has transformed human lives in treating bacterial infections before they become deadly. Bacterias have evolved mechanisms to evade the antibiotic action and developed resistance against them making them ineffective. The discovery of new antibiotics has lagged behind the development of resistance leading to threat of antimicrobial resistance (AMR) pandemic. There were 1.27 million deaths attributed to bacterial AMR in 2019 alone^1^and this is projected to rise to 10 million per year by 2050^2^ if there are no new antimicrobials or rapid diagnostics available to stem its rampant use and abuse. Currently, antimicrobial susceptibility test (AST) is examined using culture of bacterial isolates against a panel of antimicorbial compounds, a process that would take at least 48 hours. During this time, empirical prescription is common, and patients often buy antimicrobials directly from pharmacy without valid prescription which contributes towards AMR. The aim of the study was to identify trends in multidurg resistance in enterobacteriaceae and non-enterobacteriaceae family and to model and predict future multi-durg resistance trends. The AMR proportion predictions were carried out using time series analysis with existing AMR data from Venatorx Global Evaluation of Antimicrobial Resistance via Surveillance (GEARS) Program^3^. Autoregressive integrated moving average (ARIMA) is widely applied in time series analysis and aids in short term predictions based on historial AMR data^4^ to facilitate selection of appropriate antibiotics and reduce AMR outbreaks.

## Materials and Methods

We utilized GEARS (Global Evaluation of Antimicrobial Resistance via Surveillance) dataset with Data Request ID: 00009068 from AMR Vivli for our analysis (https://amr.vivli.org/). We sorted the data to include only India specific dataset.

We used the minimum inhibitory concentration (MIC) breakpoints (ug/ml) for each antibiotic based on the CLSI 2022 guidelines^5^.

The MIC predictions were carried out based on ARIMA as follows:

1. Organize the available dataset, containing MIC values for the years 2018 to 2021, into separate arrays for each antibiotic category of MIC as sensitive (S), intermediate (I), resistant (R) based on CLSI 2022 guidelines.
2. ARIMA models are applied for each category of MIC and the total. ARIMA is a time series forecasting method that takes into account the temporal nature of the data and captures patterns, trends, and seasonalities.
3. The ARIMA models are used to predict the MIC values for the year 2022, 2023, 2024 for each category, giving a single predicted value for each category.
4. The Mean Squared Error (MSE) is then computed for each ARIMA model. MSE measures the average squared difference between the predicted values and the actual values. Lower MSE values indicate better accuracy of the predictions.

Justification for ARIMA: ARIMA is used for time series forecasting when there is a temporal component in the data. In this dataset, the MIC values are recorded over different years, suggesting a temporal pattern. ARIMA models capture the underlying trends and seasonalities in the data, making them appropriate for forecasting future values. This method can account for possible changes in MIC values over time, considering the inherent variability in the data.

MSE is used for the ARIMA models to evaluate the accuracy of the predictions. It measures the average squared difference between the predicted and actual values. Lower MSE values indicate that the ARIMA models have made more accurate predictions, showing their ability to forecast future values with less error.

Overall, by employing ARIMA methods and computing MSE, we have provided a comprehensive evaluation of the predictive performance of the models, considering the data’s temporal and limited nature.

Ceftazidime, Gentamicin, Cefepime, Levofloxacin are all broad spectrum antibiotics and we sorted the clinical isolates based on their individual antibiotic susceptibility patterns into sensitive (S), intermediate (I) and resistant (R) isolates. Subsequently, we calculated percentage of S, I and R isolates for each category.

All the relevant data code used for ARIMA prediction can be found at the following GitHub repository: https://github.com/nehalkalita/AMR-analysis

## Results

| Enterobacteriaceae |  |  |  |  | % isolate(S) | % isolate(I) | % isolate(R) |
| --- | --- | --- | --- | --- | --- | --- | --- |
|  | Ceftazidime MIC (in µg/mL) |  |  |  |  |  |  |
| Year | `0 → 4` | `8` | `>= 16` | Total |  |  |  |
| 2018 | 12 | 1 | 20 | 33 | 36.4 | 3.0 | 60.6 |
| 2019 | 88 | 6 | 105 | 199 | 44.2 | 3.0 | 52.8 |
| 2020 | 33 | 4 | 36 | 73 | 45.2 | 5.5 | 49.3 |
| 2021 | 37 | 2 | 52 | 91 | 40.7 | 2.2 | 57.1 |
| Predictions ARIMA |  |  |  |  |  |  |  |
| Year | `0 → 4` | `8` | `>= 16` | Total |  |  |  |
| 2022 | 52 | 4 | 61 | 117 | 44.4 | 3.4 | 52.1 |
| 2023 | 43 | 3 | 55 | 101 | 42.6 | 3.0 | 54.5 |
| 2024 | 48 | 4 | 59 | 111 | 43.2 | 3.6 | 53.2 |
|  |  |  |  |  | 42.4 | 3.4 | 54.2 |
|  | Gentamicin MIC (in µg/mL) |  |  |  |  |  |  |
| Year | `0 → 4` | `8` | `>= 16` | Total |  |  |  |
| 2018 | 21 | 1 | 11 | 33 | 63.6 | 3.0 | 33.3 |
| 2019 | 132 | 2 | 65 | 199 | 66.3 | 1.0 | 32.7 |
| 2020 | 40 | 1 | 32 | 73 | 54.8 | 1.4 | 43.8 |
| 2021 | 52 | 4 | 35 | 91 | 57.1 | 4.4 | 38.5 |
| Predictions ARIMA |  |  |  |  |  |  |  |
| Year | `0 → 4` | `8` | `>= 16` | Total |  |  |  |
| 2022 | 74 | 2 | 42 | 118 | 62.7 | 1.7 | 35.6 |
| 2023 | 62 | 4 | 37 | 103 | 60.2 | 3.9 | 35.9 |
| 2024 | 68 | 2 | 40 | 110 | 61.8 | 1.8 | 36.4 |
|  |  |  |  |  | 60.9 | 2.5 | 36.6 |

| Enterobacteriaceae |  |  |  |  |  |  |  |
| --- | --- | --- | --- | --- | --- | --- | --- |
|  | Cefepime MIC (in µg/mL) |  |  |  |  |  |  |
| Year | `0 → 2` | `4 → 8` | `>= 16` | Total | % isolate(S) | % isolate(I) | % isolate(R) |
| 2018 | 15 | 0 | 18 | 33 | 45.5 | 0.0 | 54.5 |
| 2019 | 94 | 13 | 92 | 199 | 47.2 | 6.5 | 46.2 |
| 2020 | 34 | 6 | 33 | 73 | 46.6 | 8.2 | 45.2 |
| 2021 | 38 | 7 | 46 | 91 | 41.8 | 7.7 | 50.5 |
| Predictions ARIMA |  |  |  |  |  |  |  |
| Year | `0 → 2` | `4 → 8` | `>= 16` | Total |  |  |  |
| 2022 | 55 | 8 | 54 | 117 | 47.0 | 6.8 | 46.2 |
| 2023 | 46 | 7 | 49 | 102 | 45.1 | 6.9 | 48.0 |
| 2024 | 51 | 7 | 52 | 110 | 46.4 | 6.4 | 47.3 |
|  |  |  |  |  | 45.6 |  | 48.3 |
|  | Levofloxacin MIC (in µg/mL) |  |  |  |  |  |  |
| Year | `0 → 0.5` | `1` | `2` | Total | % isolate(S) | % isolate(I) | % isolate(R) |
| 2018 | 12 | 3 | 18 | 33 | 36.4 | 9.1 | 54.5 |
| 2019 | 84 | 8 | 107 | 199 | 42.2 | 4.0 | 53.8 |
| 2020 | 27 | 2 | 44 | 73 | 37.0 | 2.7 | 60.3 |
| 2021 | 37 | 1 | 53 | 91 | 40.7 | 1.1 | 58.2 |
| Predictions ARIMA |  |  |  |  |  |  |  |
| Year | `0 → 0.5` | `1` | `2` | Total |  |  |  |
| 2022 | 48 | 3 | 65 | 116 | 41.4 | 2.6 | 56.0 |
| 2023 | 41 | 3 | 57 | 101 | 40.6 | 3.0 | 56.4 |
| 2024 | 45 | 3 | 62 | 110 | 40.9 | 2.7 | 56.4 |
|  |  |  |  |  | 39.9 | 3.6 | 56.5 |

| Non-Enterobacteriaceae |  |  |  |  | % isolate(S) | % isolate(I) | % isolate(R) |
| --- | --- | --- | --- | --- | --- | --- | --- |
|  | Ceftazidime MIC (in µg/mL) |  |  |  |  |  |  |
| Year | `0 → 8` | `16` | `>= 32` | Total |  |  |  |
| 2018 | 10 | 1 | 4 | 15 | 66.7 | 6.7 | 26.7 |
| 2019 | 12 | 0 | 8 | 20 | 60.0 | 0.0 | 40.0 |
| 2020 | 10 | 0 | 7 | 17 | 58.8 | 0.0 | 41.2 |
| 2021 | 11 | 2 | 6 | 19 | 57.9 | 10.5 | 31.6 |
| Predictions ARIMA |  |  |  |  |  |  |  |
| Year | `0 → 8` | `16` | `>= 32` | Total |  |  |  |
| 2022 | 11 | 2 | 7 | 20 | 55.0 | 10.0 | 35.0 |
| 2023 | 11 | 2 | 6 | 19 | 57.9 | 10.5 | 31.6 |
| 2024 | 11 | 2 | 6 | 19 | 57.9 | 10.5 | 31.6 |
|  |  |  |  |  | **59.2** | **6.9** | **33.9** |
|  | Gentamicin MIC (in µg/mL) |  |  |  |  |  |  |
| Year | `0 → 4` | `8` | `>= 16` | Total |  |  |  |
| 2018 | 11 | 0 | 4 | 15 | 73.3 | 0.0 | 26.7 |
| 2019 | 19 | 0 | 1 | 20 | 95.0 | 0.0 | 5.0 |
| 2020 | 12 | 2 | 3 | 17 | 70.6 | 11.8 | 17.6 |
| 2021 | 14 | 1 | 4 | 19 | 73.7 | 5.3 | 21.1 |
| Predictions ARIMA |  |  |  |  |  |  |  |
| Year | `0 → 4` | `8` | `>= 16` | Total |  |  |  |
| 2022 | 15 | 1 | 3 | 19 | 78.9 | 5.3 | 15.8 |
| 2023 | 14 | 1 | 3 | 18 | 77.8 | 5.6 | 16.7 |
| 2024 | 14 | 1 | 3 | 18 | 77.8 | 5.6 | 16.7 |
|  |  |  |  |  | **78.2** | **4.8** | **17.1** |

| Non-Enterobacteriaceae |  |  |  |  |  |  |  |
| --- | --- | --- | --- | --- | --- | --- | --- |
|  | Cefepime MIC (in µg/mL) |  |  |  |  |  |  |
| Year | `0 → 8` | `16` | `>= 32` | Total | % isolate(S) | % isolate(I) | % isolate(R) |
| 2018 | 11 | 0 | 4 | 15 | 73.3 | 0.0 | 26.7 |
| 2019 | 12 | 1 | 7 | 20 | 60.0 | 5.0 | 35.0 |
| 2020 | 8 | 0 | 9 | 17 | 47.1 | 0.0 | 52.9 |
| 2021 | 14 | 0 | 5 | 19 | 73.7 | 0.0 | 26.3 |
| Predictions ARIMA |  |  |  |  |  |  |  |
| Year | `0 → 8` | `16` | `>= 32` | Total |  |  |  |
| 2022 | 9 | 0 | 6 | 15 | 60.0 | 0.0 | 40.0 |
| 2023 | 13 | 0 | 6 | 19 | 68.4 | 0.0 | 31.6 |
| 2024 | 10 | 0 | 6 | 16 | 62.5 | 0.0 | 37.5 |
|  |  |  |  |  | 63.6 |  | 35.7 |
|  | Levofloxacin MIC (in µg/mL) |  |  |  |  |  |  |
| Year | `0 → 1` | `2` | `>= 4` | Total | % isolate(S) | % isolate(I) | % isolate(R) |
| 2018 | 15 | 0 | 0 | 15 | 100.0 | 0.0 | 0.0 |
| 2019 | 20 | 0 | 0 | 20 | 100.0 | 0.0 | 0.0 |
| 2020 | 15 | 1 | 1 | 17 | 88.2 | 5.9 | 5.9 |
| 2021 | 19 | 0 | 0 | 19 | 100.0 | 0.0 | 0.0 |
| Predictions ARIMA |  |  |  |  |  |  |  |
| Year | `0 → 1` | `2` | `>= 4` | Total |  |  |  |
| 2022 | 15 | 0 | 0 | 15 | 100.0 | 0.0 | 0.0 |
| 2023 | 19 | 0 | 0 | 19 | 100.0 | 0.0 | 0.0 |
| 2024 | 15 | 0 | 0 | 15 | 100.0 | 0.0 | 0.0 |
|  |  |  |  |  | 98.3 | 0.8 | 0.8 |

## Conclusion

In the non-enterobacteriaceae family which includes *Pseudomonas aeruginosa, Stenotrophomonas maltophilia*, levofloxacin (98.3%) was the most susceptible broad spectrum antibiotic followed by gentamicin (78.2%), cefepime (63.6%) and ceftazidime (59.2%). On the other hand, in the enterobacteriaceae family gentamicin (60.9%) was the most susceptible antibiotic followed by cefepime (45.6%), ceftazidime (42.4%) and levofloxacin (39.9%). On the contrary, levofloxacin was the most resistant amongst the four antibiotics (56.5%) in the enterobacteriaceae family. Ceftazidime, Levofloxacin are among the Indian Council of Medical Research (ICMR) list of ‘alert antimicrobial agents’^6^ responsible for the major antimicrobial resistance patterns due to their improper prescription. Based on the future trends, hospitals and clinicans can exercise care while prescribing these antibiotics. Previous studies, have shown that levofloxacin MICs within the historically susceptible range of 1 or 2 μg/ml (pre-2019 CLSI breakpoints) in patients infected with *Enterobacterales* were associated with worse clinical outcome for mortality than MICs of ≤0.5 μg/ml^7^. Previous study from Indonesia has shown levofloxacin to be highly sensitive against enterobacterial and non-enterobacterial uropathogens^8^ which is in line with our work.

## Data Availability

All data produced in the present study are available upon reasonable request to the authors. All the relevant data code used for ARIMA prediction can be found at the following GitHub repository: https://github.com/nehalkalita/AMR-analysis

## Acknowledgements

This publication is based on research using Global Evaluation of Antimicrobial Resistance via Surveillance (GEARS) Program data from Venatorx, obtained through https://amr.vivli.org. RN is founder of Diagopreutic pvt ltd, involved in rapid diagnosis kit development. RN is also founder of Manhar foundation, a non-profit group advocating cost effective and rapid diagnosis for all.

